# Multisystem inflammatory syndrome in European White children – study of 274 cases

**DOI:** 10.1101/2021.03.30.21254584

**Authors:** Kamila Maria Ludwikowska, Magdalena Okarska-Napierała, Natalia Dudek, Paweł Tracewski, Jacek Kusa, Krzysztof Piwoński, Aneta Afelt, Dominik Cysewski, Mateusz Biela, Bożena Werner, Teresa Jackowska, Catherine Suski, Miron Bartosz Kursa, Ernest Kuchar, Leszek Szenborn

## Abstract

**Background:** Despite the growing literature on multisystem inflammatory syndrome in children (MIS-C), the data in European White population is limited. Our aim was to capture MIS-C emergence in Poland (central Europe) and to describe its characteristics with a focus on severity determinants.

**Methods:** Patients who met the MIS-C definition (fever, multiorgan failure, inflammation, and proven SARS-CoV-2 infection or contact) were reported retrospectively and prospectively in an online survey. Study definitions fulfilment was automatically evaluated by a dedicated software. For the assessment of univariate relationships, either directed or divided by sex, age, or disease severity, we used the test for two categorical variables and the Kruskal-Wallis test for categorical-continuous variable pairs.

**Findings:** The analysis involved 274 children, 62.8% boys, median age 8.8 years. Besides one Asian, all were European White. Merely 23 (8.4%) required paediatric intensive care treatment (PICU). They were older (11.2 vs. 8.4 years), and at hospital admission had higher respiratory rate (30 v. 20/minute), lower systolic blood pressure (89 vs. 100 mmHg), prolonged capillary refill time (40% vs. 11%), and decreased consciousness (22% vs. 5%).

Teenage boys had more common cardiac involvement (fraction 25.9% vs. 14.7%) and macrophage activation syndrome (31.0% vs. 15.2%) than others. Boys were also more often hospitalised in PICU with age (from median 11.2 years to 9.1).

**Interpretation:** The severity of MIS-C is not as uniform as it seemed, ethnicity and sex may affect MIS-C phenotype. Management might not be universally applicable and should rather be adjusted to the specific population.

**Funding:** PSP: 501-D402-20-0006100

## Introduction

Paediatric inflammatory multisystem syndrome temporally associated with severe acute respiratory syndrome coronavirus 2 (SARS-CoV-2) or multisystem inflammatory syndrome in children (herein referred to as MIS-C), is a new paediatric entity which has emerged in countries particularly hit by coronavirus disease 2019 (COVID-19) pandemic.^1-9^ Despite similarities to other inflammatory conditions, e.g. Kawasaki disease (KD), macrophage activation syndrome (MAS), or toxic shock syndrome, MIS-C has its distinct features.^8,9^ MIS-C is characterised by a sudden onset of rapidly progressing multisystem inflammation which particularly affects the cardiovascular system, resulting in cardiac dysfunction and shock. Milder forms of fever and inflammation were also described, however in the largest published cohorts majority of patients necessitated treatment in a paediatric intensive care unit (PICU).^4-9^

In order to perceive the emergence of MIS-C in our country, we have launched a national surveillance of inflammatory disorders in children (MultiOrgan Inflammatory Syndromes COVID-19 Related Study, MOIS-CoR). Following COVID-19 second wave in Autumn 2020, a rise in MIS-C prevalence has emerged in Poland. Here we report the results of the survey, revealing a consistent picture of MIS-C in our population, yet with some unique local characteristics.

## Material and Methods

### Data sources

The surveillance was launched on 25th May 2020. The inclusion criteria are presented in Appendix (p 2). Ethical approval was obtained from the Bioethics Committee at Wroclaw Medical University (CWN UMW BW: 313/2020). Waiver of informed consent was obtained with only deidentified data transmitted and analysed.

Children aged 0-18 years old with inflammatory conditions were voluntarily reported from 42 cities from all over the country (appendix p 8). Anonymised patients’ data were retrospectively and prospectively extracted from health records and collected through an online questionnaire developed for that purpose. Demographic data, clinical characteristics, laboratory parameters, cardiovascular evaluation results, treatment and outcome data were collected. Vital signs and laboratory parameters were obtained at admission and at their respective peaks. Here we report the data covering period between 4th March 2020 (when the first COVID-19 case was confirmed in Poland) and 20th February 2021. The analysis was data-driven. Nine of the presented cases were included in our previous cursory report.^10^

### MIS-C Case Definition

For the sake of this study, we adopted World Health Organization (WHO) MIS-C case definition^3^, as follows:

- children 0-18 years old with fever lasting at least three days AND
- at least two of the following:
  - rash or bilateral conjunctivitis, or mucocutaneous inflammation signs
  - hypotension defined by a minimal systolic blood pressure (sBP) below 70+2×age (in years) mmHg or below 90 mmHg for children older than ten years
  - features of myocardial dysfunction, pericarditis, or coronary artery abnormality (CAA), based on echocardiographic findings or elevated B-type natriuretic peptide (BNP)/N-terminal-pro-BNP (NT-proBNP), or troponin
  - evidence of coagulopathy (by international normalised ratio [INR] >1.1, activated partial thromboplastin time >40 sec or D-dimer >500 µg/mL)
  - acute gastrointestinal problems.
  AND
- elevated inflammatory markers: ESR ≥40 mm/hr, C-reactive protein (CRP) ≥30 mg/L, or procalcitonin ≥0.5 ng/mL AND
- no other apparent microbial cause AND
- evidence of COVID-19 (positive real-time polymerase chain reaction (RT-PCR), antigen test, or serology), or personal history of COVID-19 or contact with a proven COVID-19 case.

### Standardised Study Definitions

#### Laboratory Abnormalities

We defined lymphopenia as lymphocyte count <1.5 ×10^9^/L, anaemia according to age-related norms, thrombocytopenia as platelets <150 ×10^9^/L, elevated alanine transaminase as ≥40 U/L, hyponatremia as serum sodium <135 mmol/L, hypoalbuminemia as serum albumin <3.5 g/dL, elevated BNP/NT-proBNP as >150 ng/mL. A threshold of 50 ng/L defined elevated both T and I troponin. Renal dysfunction was defined by estimated glomerular filtration rate (eGFR) <90 ml/min/1.73 m^2^, calculated using the revised Schwartz formula.

#### Echocardiographic Abnormalities

Echocardiography results were categorised based on descriptive results and left ventricular ejection fraction (EF) and coronary artery measurements whenever available. Heart dysfunction was defined as EF <55% and severe heart dysfunction as EF <35%. Coronary artery Z-score was calculated using Dallaire equation or Boston Children’s Hospital Z-score calculator, depending on the body surface area. Dilation was defined by Z-score between 2 to 2.5, while aneurysm by Z-score ≥2.5.^11^ The worst available EF and the largest coronary Z-scores were included. The echography results were assessed by two independent cardiologists.

#### Clinical Definitions

Diagnostic criteria of KD in its typical and atypical (aKD) form were adapted from American Heart Association (AHA) guidelines.^11^ MAS was diagnosed based on Paediatric Rheumatology International Trials Organization criteria (consult appendix p 2).^12^

The level of consciousness was evaluated using the AVPU scale.

Nutritional status was assessed using the body-mass index (BMI), converted into Z-scores based on the WHO reference standards for children younger than 5 years^13^ and national reference standards for older children.^14^

#### Statistical Methods

We describe variables in relation to the sum of cases for which the variable was recorded. We assumed missing values to be distributed randomly and independently from the data, and propagated them through aforementioned clinical definitions according to łukasiewicz logic. Fulfilment of all aforementioned study definitions was automatically evaluated by a dedicated software.

For the assessment of univariate relationships, either directed or divided by sex or age group, we used Cochran-Mantel-Haenszel test for two categorical variables and Kruskal-Wallis test for categorical-continuous variable pairs. Confidence intervals for incidence values were estimated according to Clopper-Pearson method. Significance level of 0.05 and two-sided testing was employed. All statistical analyses were performed in R, version 4.0.3 (R Foundation for Statistical Computing), using coin and GenBinomApps packages. MIS-C incidence was estimated based on the demographic data published by Polish Central Statistical Office.

Due to an exploratory nature of this study, we have not adjusted p-values for multiple comparisons.

## Results

### Study Group

As of 20th February 2021, 399 children have been registered to the surveillance, 342 of whom fulfilled the inclusion criteria and 274 fulfilled MIS-C diagnostic criteria (Figure 1 and appendix p 4). The following results apply only to the cohort of 274 children, aged from 7 months to 17.6 years, with MIS-C.

**Figure 1.**
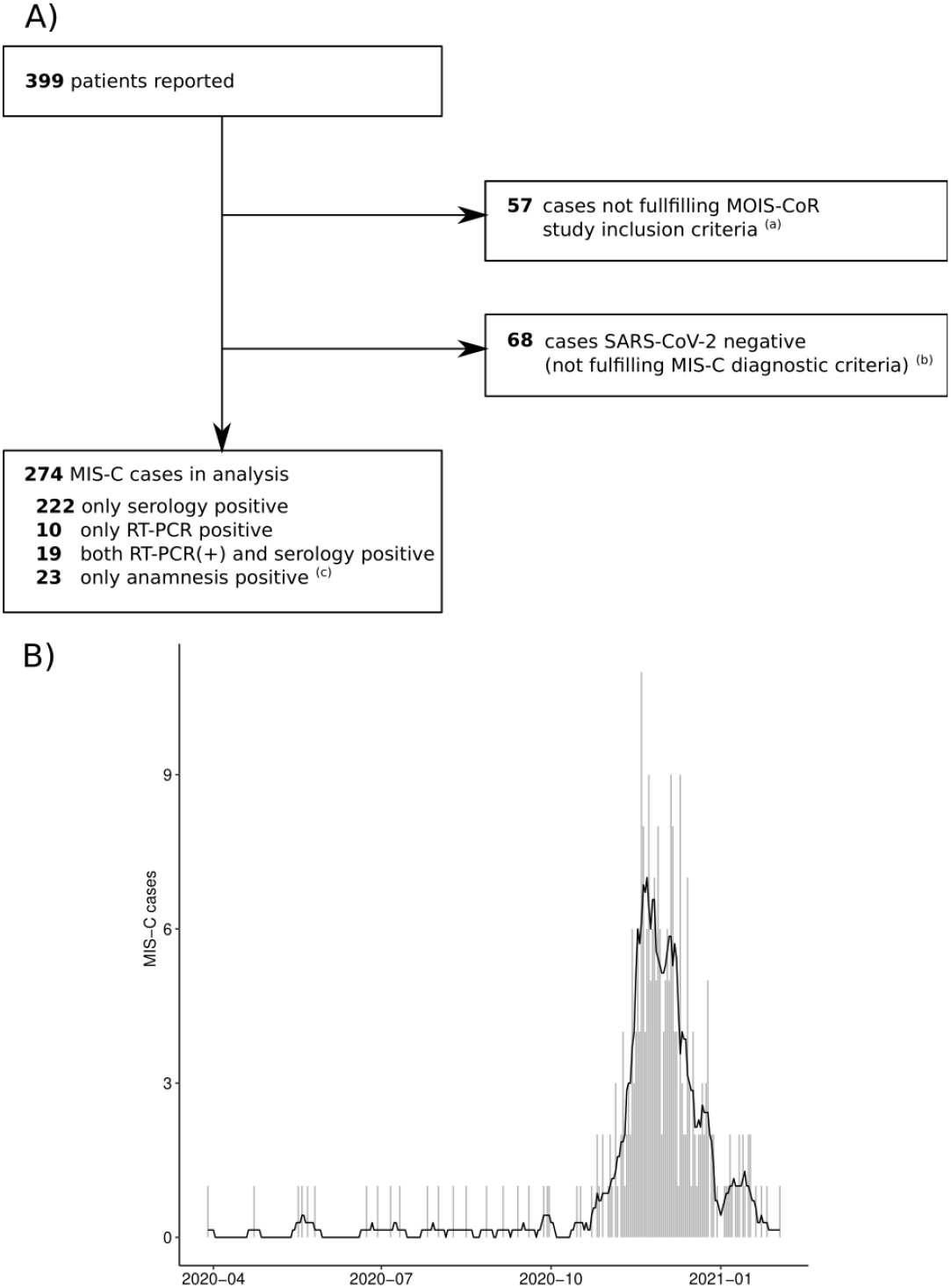
Eligibility Flowchart of patients reported in MultiOrgan inflammatory Syndromes COVID-19 Related Study (MOIS-CoR) and temporal distribution of MIS-C cases in Poland A) Eligibility Flowchart of patients reported in MultiOrgan inflammatory Syndromes COVID-19 Related Study (MOIS-CoR), 4th March 2020 to 20th February 2021 Abbreviations: MIS-C, multisystem inflammatory syndrome in children; MOIS-CoR, MultiOrgan inflammatory Syndromes COVID-19 Related Study; RT-PCR, real-time transcription polymerase chain reaction; SARS-CoV-2, severe acute respiratory syndrome coronavirus 2 ^a^ Study inclusion criteria are presented in STable 1. ^b^ MIS-C criteria were based on the World Health Organization definition, precise criteria are presented in Materials and Methods section. ^c^ Anamnesis encompassed history of previous confirmed SARS-CoV-2 infection or contact with a proven coronavirus disease 2019 (COVID-19) B) Temporal distribution of MIS-C cases in Poland, 4th March 2020 to 20th February 2021 Bars indicate daily number of cases and line indicates weekly average number of cases

Demographic characteristics of the MIS-C cohort are presented in Table 1. All but one Asian child were of European White ethnicity, while 172 (62.8% [95%CI 56.8%-68.5%]) were male. A minority of children had any comorbidities, with obesity (6.7% [95%CI 3.9%-10.6%]), asthma (4.1% [95%CI 2.1%-7.3%]) and neurological disorders (2.3% [95%CI 0.8%-4.9%]) being the most prevalent.

**Table 1.**
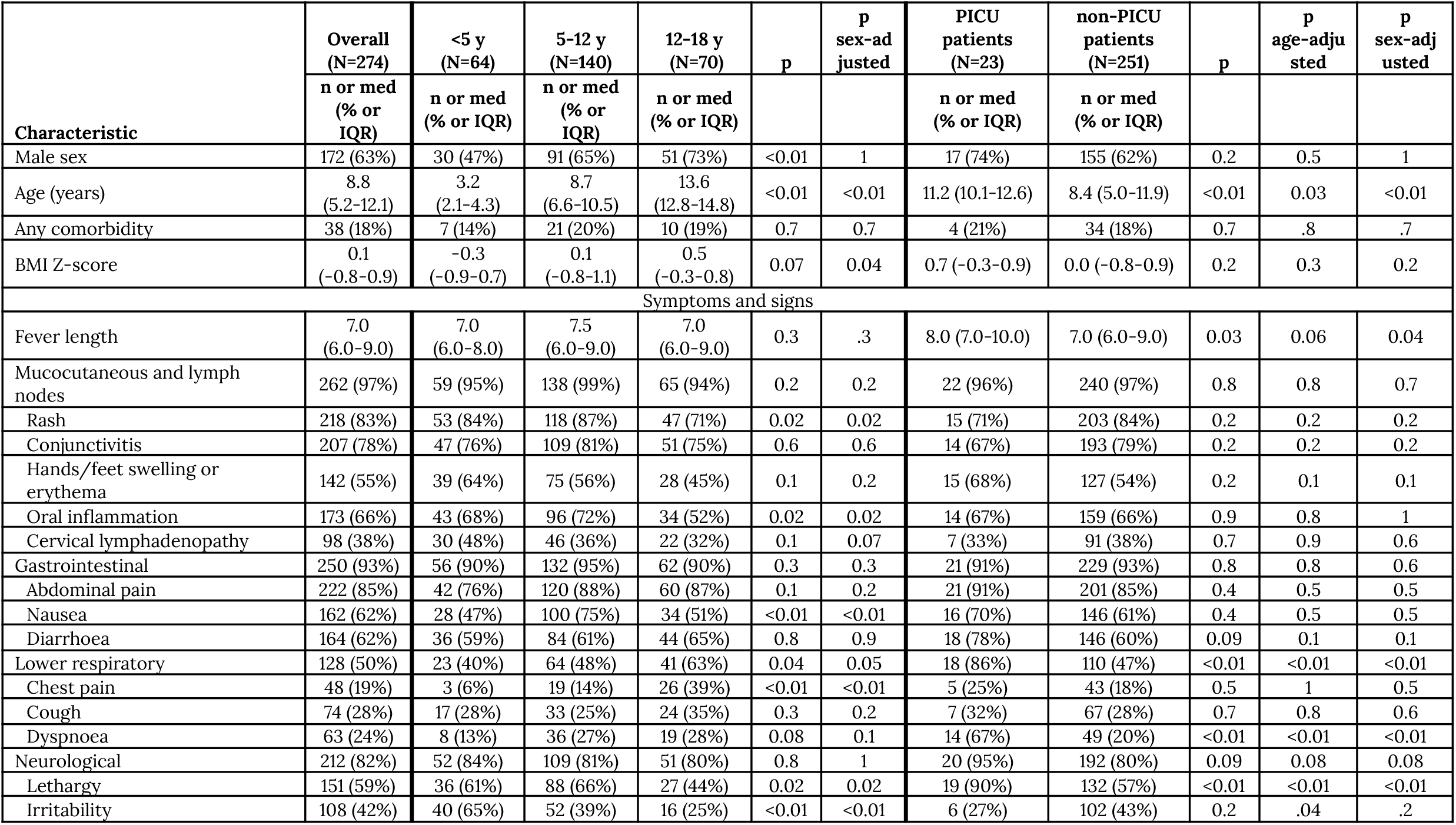
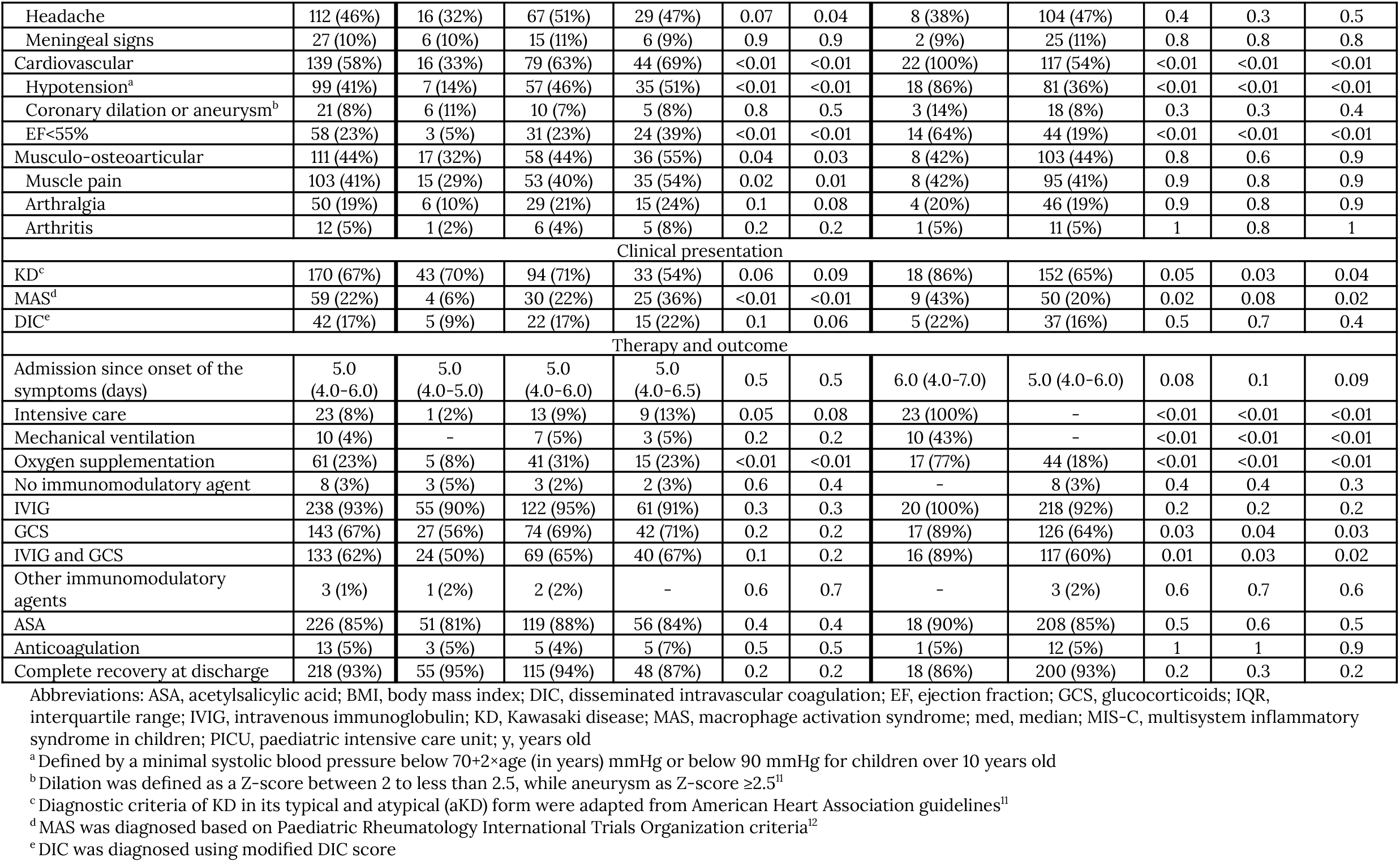
Demographic and clinical characteristics, management and outcome of MIS-C cohort

### Clinical Presentation

The median time between first symptoms and hospital admission was five days, and the median fever length was seven days. The vital signs at admission and at their respective peaks are presented in Table 2. Complete clinical characteristics of children with MIS-C are presented in Table 1 and appendix (p 5).

**Table 2.**
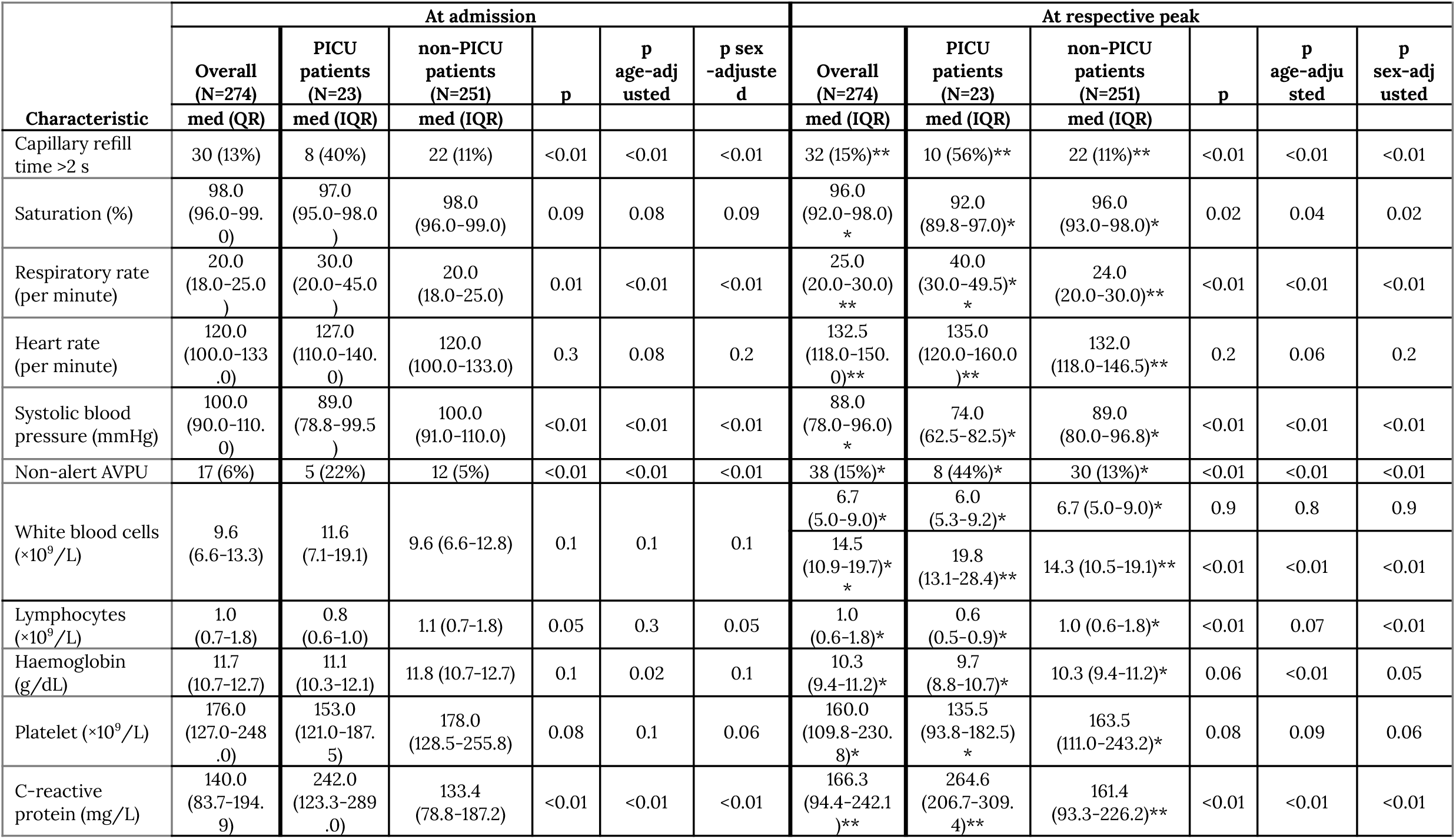
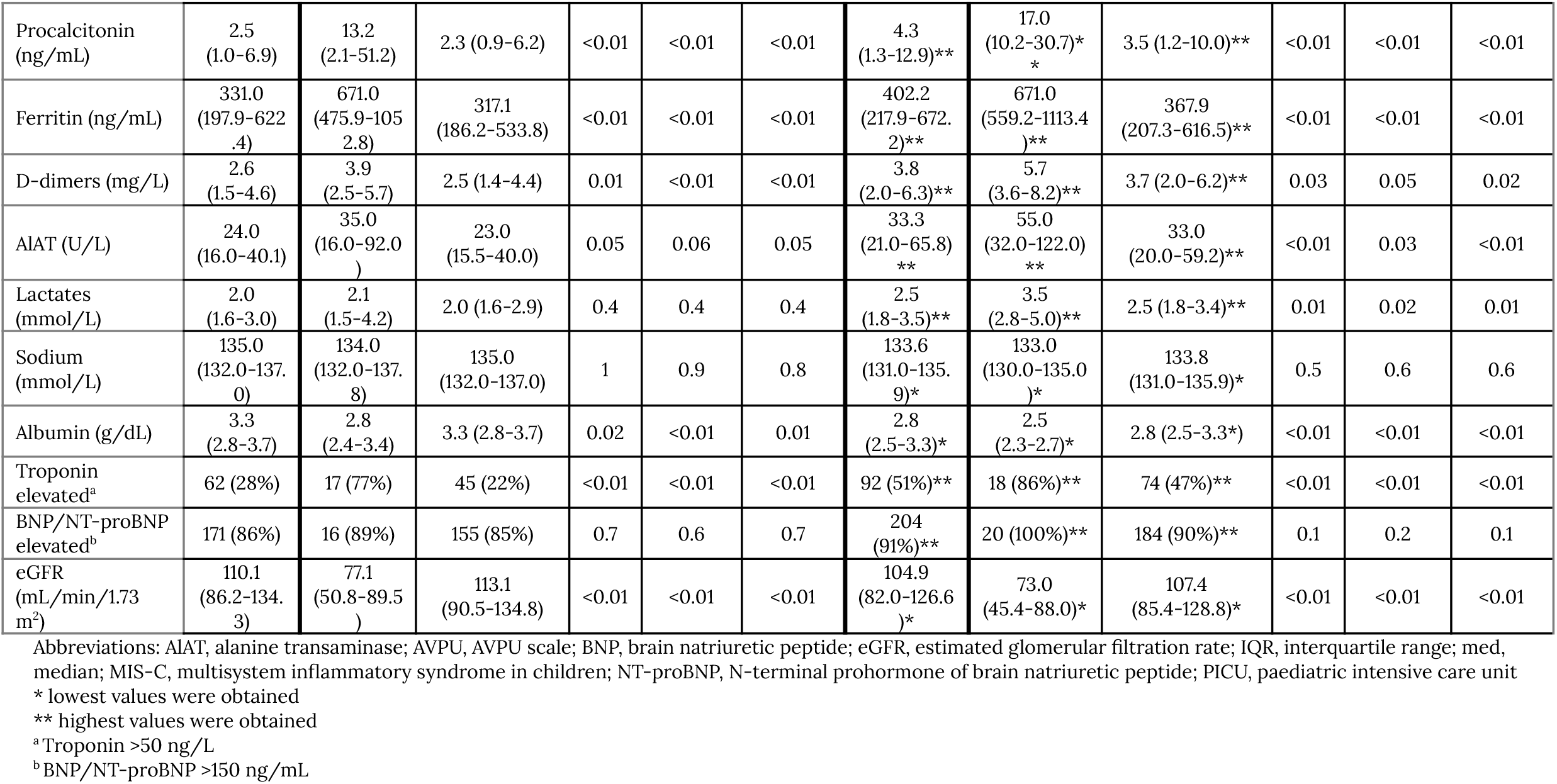
Vital signs and laboratory results of MIS-C cohort at admission and at respective peaks.

Mucocutaneous and lymph node involvement was observed in 95.6% (95%CI 92.4%-97.7%) of children, and 66.7% (95%CI 60.5%-72.4%) fulfilled AHA KD/aKD diagnostic criteria, irrespective of age. Rash was less common in children >12 years of age (71.2% [95%CI 58.7%-81.7%] vs. 86.4% [95%CI 80.8%-90.8%], p<0.01), whereas conjunctival injection, distal extremity changes and cervical lymphadenopathy were equally prevalent across age groups.

The second most common group of associating symptoms was gastrointestinal (92.6% [95%CI 88.8%-95.4%]), with no significant differences across age groups, except for nausea and vomiting, which were more prevalent in children 5-12 years of age (74.6% [95%CI 66.4%-81.7%] vs. 48.8% [95%CI 39.9%-57.8%], p<0.01). Ten children underwent abdominal surgery due to acute abdominal symptoms.

Hypotension was present in 30/213 (14.1% [95%CI 9.7%-19.5%]) of patients at admission, while 99/243 (40.7% [95%CI 34.5%-47.2%]) developed it at some point during hospitalization. At least one echocardiogram was reported for 255 children, of whom 85 (33.3% [95%CI 27.6% to 39.5%]) had any of the following changes: decreased EF, CAA or pericardial effusion. EF <55% was reported in 58/255 (22.7% [95%CI 17.7%-28.4%]) of children (four had EF <35%), and heart dysfunction overall was significantly more prevalent with age (from median 11.0 years [IQR 8.8-13.6] to 7.9 [4.9-11.3], p<0.01). CAA developed in 21/255 (8.2% [95%CI 5.2%-12.3%]) of patients irrespectively of age or KD/aKD phenotype. In particular, eight children developed coronary artery aneurysms; three of them resolved in the follow up before discharge. Pericardial effusion was present in 24/255 (9.4% [95%CI 6.1%-13.7%]) of reports.

Neurological symptoms included lethargy (59.4% [95%CI 53.1%-65.5%]), irritability (41.7% [95%CI 35.6%-48.0%]), headache (46.1% [95%CI 39.7%-52.6%]) and photophobia (11.0% [95%CI 7.4%-15.5%]). Children >12 years old less frequently presented lethargy (44.3% [95%CI 31.5%-57.6%] vs. 64.2% [95%CI 57.0%-71.0%], p<0.01) and irritability (25.0% [95%CI 15.0%-37.4%] vs. 47.2% [95%CI 40.0%-54.4%], p<0.01). Meningeal signs were observed in 27/260 (10.4%, [95%CI 7.0%-14.7%]) of cases, irrespective of age, and six patients had aseptic meningitis.

Respiratory symptoms included sore throat (34.0% [95%CI 28.2%-40.2%]), cough (28.1% [95%CI 22.8%-34.0%]) and dyspnoea (24.1% [95%CI 19.1%-29.8%]), and they were uniformly prevalent across age groups. Patients >12 years old more often complained of chest pain (39.4% [95%CI 27.6%-52.2%] vs. 11.8% [95%CI 7.5%-17.3%], p<0.01) and muscle pain (53.8% [95%CI 41.0%-66.3%] vs. 36.6% [95%CI 29.6%-43.9%], p=0.01).

Dysuria was reported in 40/257 (15.6% [95%CI 11.4%-20.6%]) of children, while sterile leukocyturia in 48/247 (19.4% [95%CI 14.7%-24.9%]).

### Laboratory Results

Laboratory parameters at admission and at respective peaks are summarised in Table 2. Lymphopenia was present in 195/269 (72.5% [95%CI 66.7%-77.7%]) of children, and 148/269 (55.0% [95%CI 48.9%-61.1%]) developed lymphopenia of <1.0 ×10^9^/L. Anaemia was present in 254/273 (93.0% [95%CI 89.3%-95.8%]) of patients. Thrombocytopenia occurred in 131/273 (48.0% [95%CI 41.9%-54.1%]) of children, and 31/273 (11.4% [95%CI 7.8%-15.7%]) had a platelet count below 80×10^9^/L.

Following MIS-C case definition, all children had increased inflammatory markers: 158/266 (59.4% [95%CI 53.2%-65.4%]) had procalcitonin >2.5 ng/mL and 157/273 (57.5% [95%CI 51.4%-63.4%]) had CRP >150 mg/L. Hypoalbuminemia was present in 219/260 (84.2% [95%CI 79.2%-88.4%]) of patients, and in 71/260 (27.3% [95%CI 22.0%-33.2%]) albumin level fell under 2.5 g/dL. Hyponatremia was found in 203/272 (74.6% [95%CI 69.0%-79.7%]) of patients, and in 9/272 (3.3% [95%CI 1.5%-6.2%]) it was <125 mmol/L.

Finally, 62/222 (27.9% [95%CI 22.1%-34.3%]) of patients had elevated troponin at admission, and this ratio increased to 51.4% (95%CI 43.8%-58.9%) when assessed later in course of the disease. Troponin levels were more commonly elevated in older children, both at admission (from median 11.4 [IQR 9.0-13.5] to 7.9 [4.9-10.7], p<0.01) and at its peak (from median 11.2 [IQR 8.7-13.4] to 8.5 [5.0-11.9], p<0.01).

### Sex-dependent Clinical and Laboratory Characteristics

Male patients were diagnosed with MIS-C more often than expected from demographic structure, but only in the older age bracket (Figure 2). We have identified some characteristics which corresponded with this discrepancy (Figure 3, see also Table 1 and Table 2 for sex-adjusted p-value).

**Figure 2.**
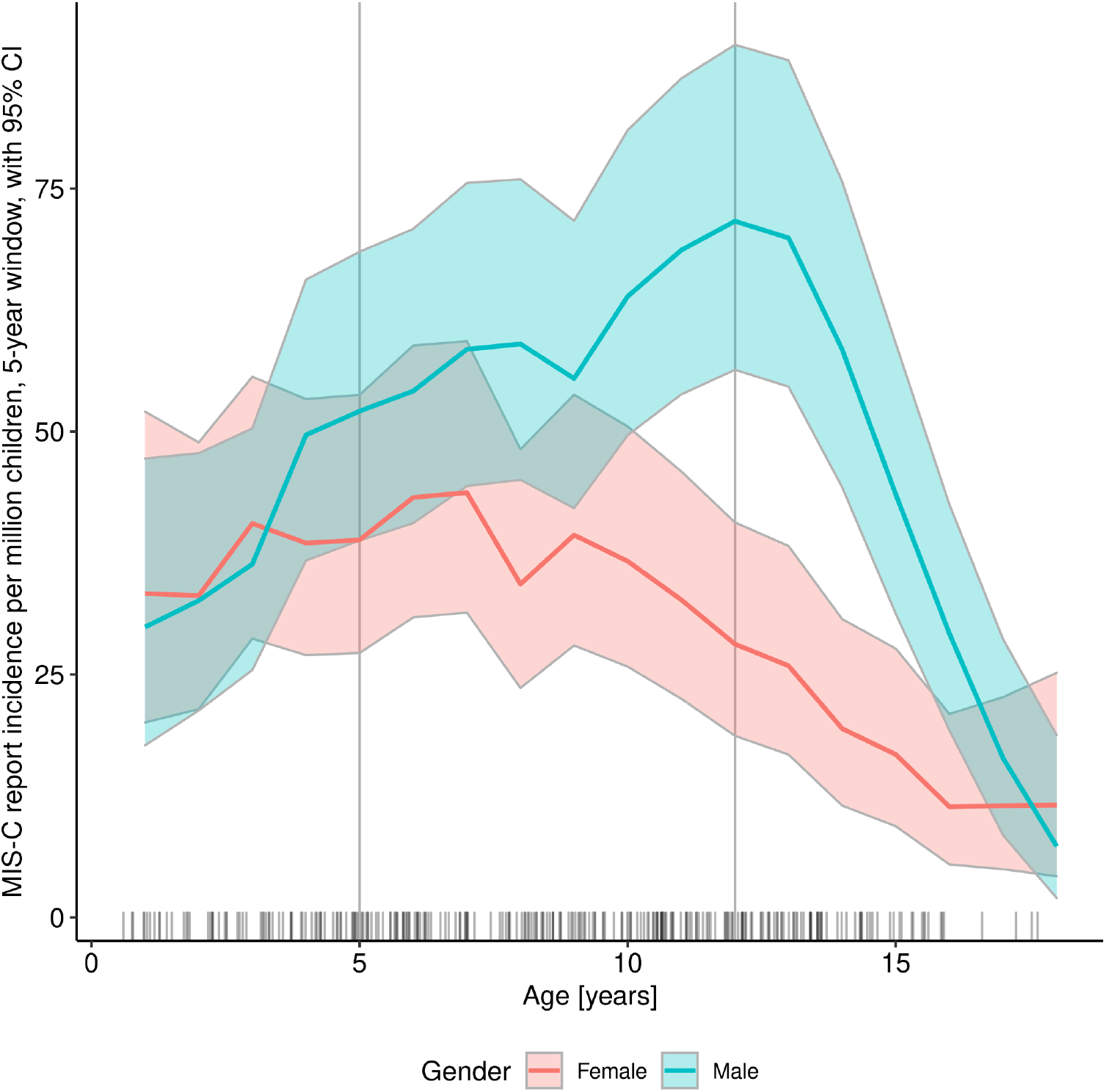
Incidence of reported MIS-C cases within the Polish population of children aged 0-18 years, according to age and sex Abbreviations: MIS-C, multisystem inflammatory syndrome in children

**Figure 3.**
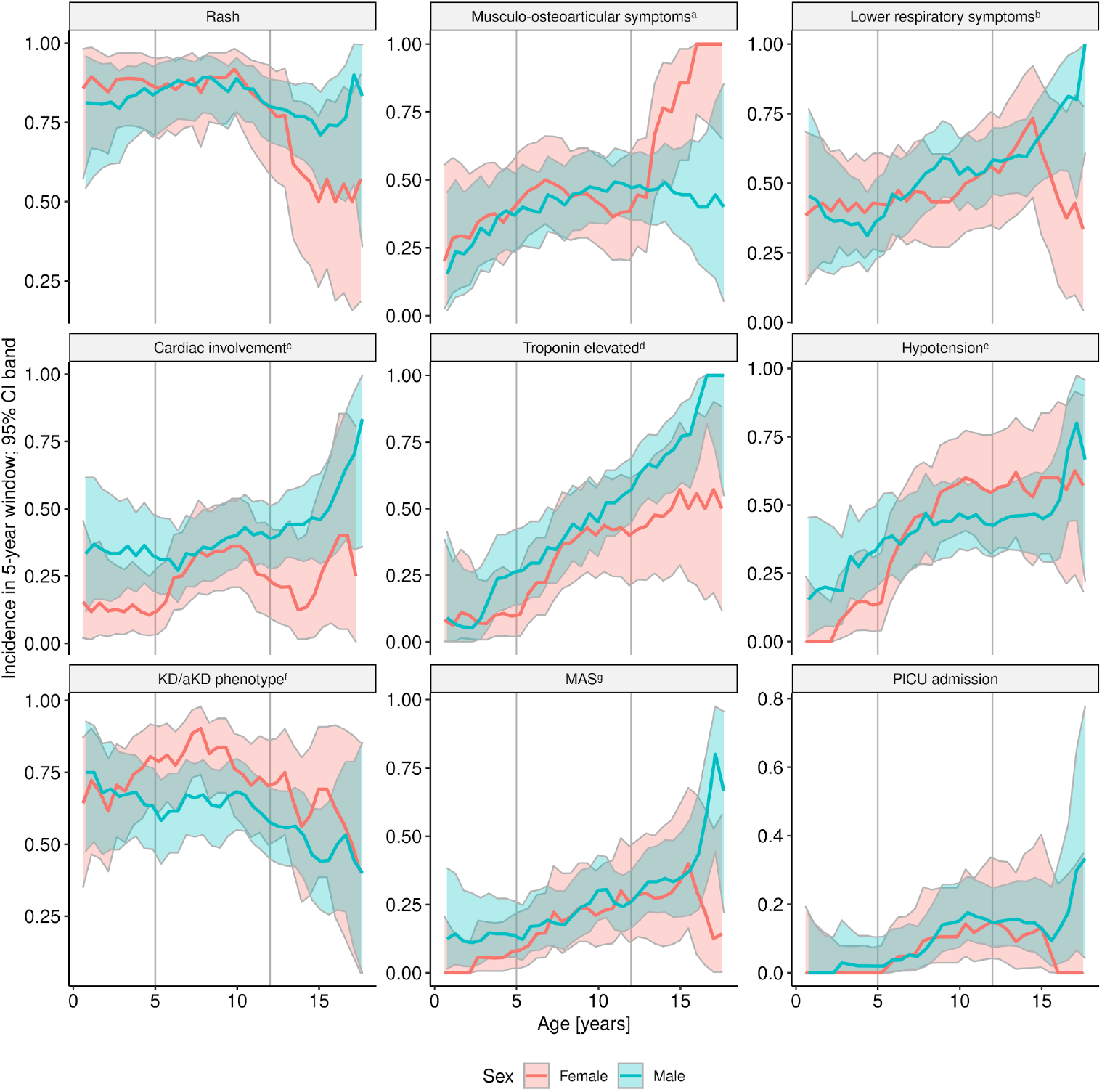
Incidence of the selected events within the study group, according to age and sex Abbreviations: KD/aKD, Kawasaki disease/atypical Kawasaki disease; MAS, macrophage activation syndrome; MIS-C, multisystem inflammatory syndrome in children; PICU, paediatric intensive care unit ^a^ Musculo-osteoarticular symptoms encompassed: muscle pain, arthralgia, or arthritis ^b^ Lower respiratory symptoms encompassed: chest pain, cough, or dyspnea ^c^ Cardiac involvement encompassed left ventricular ejection fraction <55%, or coronary artery abnormalities (dilation or aneurysm) or pericardial effusion ^d^ The highest available result of troponin concentration was included in this analysis ^e^ defined by a minimal systolic blood pressure below 70+2×age (in years) mmHg or below 90 mmHg for children over 10 years old ^f^ Diagnostic criteria of KD/aKD were adapted from American Heart Association guidelines^11^ ^g^ MAS was diagnosed based on Paediatric Rheumatology International Trials Organization criteria^12^

Teenage boys over 12 years old more prevalently had cardiac involvement (25.9% [95%CI 17.0%-36.5%] vs. 14.7% [95%CI 9.7%-20.9%], p=0.03) and fulfilled MAS diagnostic criteria more often (31.0% [95%CI 19.5%-45.5%] vs. 15.2% [95%CI 10.6%-20.7%], p<0.01). Boys were also more often hospitalised in PICU with age (from median 11.2 [IQR 10.3-12.6] to 9.1 [5.7-12.3], p=0.02), while there was no such trend for girls.

On the other hand, teenage girls more frequently presented osteoarticular and muscular symptoms (12.6% [95%CI 7.1%-20.3%] vs. 2.9% [95%CI 0.8%-7.2%], p<0.01), but less frequently rash (5.0% [95%CI 2.5%-8.8%] vs. 15.2% [95%CI 6.3%-28.9%], p=0.01). KD/aKD phenotype prevalence did not differ between reported girls and boys of neither age.

### Treatment and Outcome

Treatment is presented in Table 1. PICU treatment was required in 23/274 (8.4% [95%CI 5.4%-12.3%]) of children; ten of whom were mechanically ventilated. There were no children treated with extracorporeal membrane oxygenation (ECMO), neither with renal replacement therapy. Two deaths were reported: one in severely immunocompromised child, and one in previously healthy teenager with fulminant multiorgan dysfunction, both with positive RT-PCR test result for SARS-CoV-2. In either case, it was impossible to determine whether the cause of death was a cytokine storm due to COVID-19 or MIS-C; they fulfilled the MIS-C criteria though, and hence were included in this analysis.

Children who were hospitalised in PICU were significantly older (from median 11.2 years [IQR 10.1-12.6] to 8.4 [5.0-11.9], p<0.01). The PICU admission rate did not significantly differ for patients with comorbidities, including obesity (Table 1 and appendix p 5). The median time of hospital admission since the first symptoms was five days and it was not significantly different in children admitted to PICU versus the others. At admission, children who necessitated intensive care, had higher respiratory rate (from median 30.0/minute [IQR 20.0-45.0] to 20.0 [18.0-25.0], p=0.01) and lower sBP (from median 89.0 mmHg [IQR 78.8-99.5] to 100.0 [91.0-110.0], p<0.01), despite their higher age. They were also more likely to have prolonged capillary refill time (CRT) (40.0% [95%CI 19.1%-63.9%] vs. 10.8% [95%CI 6.9%-15.9%], p<0.01) and AVPU scale score below A (21.7% [95%CI 7.5%-43.7%] vs. 5.0% [95%CI 2.6%-8.6%], p<0.01). PICU-hospitalised children had also significantly higher levels of CRP (from median 242.0 mg/dL [IQR 123.3-289.0] to 133.4 [78.8-187.2], p<0.01), procalcitonin (from median 13.2 ng/mL [IQR 2.1-51.2] to 2.3 [0.9-6.2], p<0.01), ferritin (from median 671.0 μg/L [IQR 475.9-1052.8] to 317.1 [186.2-533.8], p<0.01) and D-dimers (from median 3.9 μg/mL [IQR 2.5-5.7] to 2.5 [1.4-4.4], p=0.01). In the later course, children in PICU had significantly more prevalent lymphopenia (81.0% [95%CI 58.1%-94.6%] vs. 47.0% [95%CI 40.5%-53.6%], p<0.01), and hypoalbuminemia (100.0% [95%CI 84.7%-100.0%] vs. 81.0% [95%CI 75.0%-86.0%], p=0.04).

## Discussion

In this study we have described the largest published so far cohort of European White children with confirmed MIS-C. Despite being initially reported as Kawasaki-like disease, MIS-C appeared to be a distinct entity soon after.^8,9^ Children in our cohort fulfilled KD/aKD diagnostic criteria more frequently than in other reports, but they concomitantly presented with unique features typical for MIS-C. On the other hand, in contrast to Western European and the United States (US) reports, the severity of the disease appeared substantially milder, as expressed by only 8.4% of patients hospitalised in PICU and two deaths. Moreover, the clinical and laboratory picture was more uniform across age groups in our cohort.^4-6,8,9^ These discrepancies might be related to more homogenous genetic and racial characteristics of the Polish population. Children of Black, South Asian and/or Hispanic ethnicity predominated in the clusters of MIS-C reported thus far.^4-6,8,9^ Non-Hispanic White children comprised only 13-30% of cases in the most numerous MIS-C cohorts and systematic reviews.^4-6,8,9^ Importantly, the proportion of Black and Hispanic ethnicity in MIS-C groups was significantly higher than in local societies.^7,15^ It is not clear whether this overrepresentation of ethnic minorities and underrepresentation of non-Hispanic White children among MIS-C patients reflect genetic predisposition or higher exposure to SARS-CoV-2, as COVID-19 disproportionately affected Hispanic and Black subpopulations.^15,16^

Two hundred seventy-four MIS-C cases captured in Poland with a 7.31 million children population as compared to 2060 cases reported in the USA with 74 million children at the same time,^1,17^ suggest that MIS-C prevalence in our country could have reached a level comparable to the US, despite specific, homogenous racial background.

Another issue is whether race/ethnicity is associated with the severity of the disease. Some authors suggest that children of Black ethnicity present a more severe clinical course of MIS-C,^4,6^ whereas others^18^ argue against it. Our cohort was almost exclusively composed of European White children which is a major demographic characteristic distinguishing it from other MIS-C cohorts. This should be considered as a possible explanation of milder clinical presentation with favourable outcome, however, this conclusion should be treated with caution and requires further analysis.

Another feature of our cohort was the relatively small proportion of obese children as compared to majority of reports from other countries (6.7% vs. 18-26%).^4,6,9^ This could have possibly resulted from lower obesity prevalence among children in Poland (up to 13%).^19^ Interestingly, in the US, obesity is more prevalent in Black (up to 22%) and Hispanic (up to 26%) children as opposed to non-Hispanic White (up to 14.1%).^20^ It is unknown whether obesity is a risk factor for developing MIS-C nor if it is connected to its severity. In our study, we found no association between BMI Z-score or obesity and severity of the disease.

Polish recommendations for MIS-C treatment are similar to those from the USA or the United Kingdom.^21,22^ Treatment used in our cohort did not differ substantially from treatment reported by other authors – most children received IVIG and a large proportion also got steroids. Fewer children required more than two different immunomodulatory agents.^4-6,8,9^ Moreover, the median day of hospital admission since the first symptoms was similar to other reports.^6,23^ Hence, the therapeutic approach is an unlikely factor of a more favourable outcome in Polish children with MIS-C.

Despite relatively uniform clinical presentation in terms of mucocutaneous, gastrointestinal, or respiratory manifestations across the age groups, cardiovascular involvement significantly increased with age, which is in line with Dufort and Abrams findings.^4,23^ Laboratory markers of the heart injury were elevated in the majority of patients, whereas hypotension was present in 40.7% and decreased EF – in 22.6% of patients.

Cardiac involvement is a major factor determining MIS-C severity, however data about cardiovascular complications are inconsistent. This is partially due to varying (sometimes unspecified) definitions used by different authors,^4,6,23,24^ and varying inclusion criteria – either broader than WHO MIS-C case definition,^24^ or narrowed only to the most severe cases.^7^ Decreased sBP, occasionally defined as a shock, was reported in 36-86% of patients with MIS-C, whereas decreased left ventricular function – 20-45%.^4-8,24^ Our findings place Polish children with MIS-C within the “milder end” of the acute cardiovascular complications spectrum described above.

Similarly, the prevalence of coronary artery involvement in MIS-C is debatable. Undoubtedly aneurysms may complicate the disease, cases of giant aneurysms have been described,^24^ but the true prevalence of aneurysms is unknown. CAA prevalence may be overestimated, as coronary dilation may be a result of an acute febrile condition or myocarditis.^25^ Both coronary artery aneurysms and dilations were less prevalent in our group than in other reports.^4-6,8,9,24^

Interestingly, we have observed that some MIS-C features differed between girls and boys in adolescence and, to our knowledge, this is the first report of sex-related discrepancies in MIS-C presentation. Teenage boys had more frequent cardiac involvement, MAS and PICU hospitalization rate. The more severe course of COVID-19 in adult males is well established.^26^ While some authors postulate that it is linked with genetic and immunological background,^27^ others suggest that sex hormones may play a role.^28,29^ COVID-19 and MIS-C are separate entities, but share some similarities being hyperinflammatory conditions. In our cohort, the distinction between children of different sex appeared at pubertal age, which might support the hormonal theory.

Finally, we aimed to identify clinical and laboratory features specific to patients who required intensive care. Similarly to previous reports, the only demographic characteristic associated with PICU admission was older age.^4,6^ The median time of hospital admission since the first symptoms did not differ significantly for PICU patients. They could be distinguished by their vital signs at hospital admission: decreased level of consciousness, longer CRT, higher respiratory rate and lower sBP. In concordance with other reports, the severity of disease was correlated with the particularly high inflammatory markers.^6,8^ The involvement of the laboratory evaluation at admission is one of strengths of our study; we found that initial values of inflammatory markers, D-dimers, albumin, eGFR, and markers of the heart injury correlated with the progression to severe disease.

## Limitations

The study relied on voluntary participation, hence a number of MIS-C cases might have been missed or biased by non-random sampling. Some patients meeting the MIS-C criteria may have been misclassified, e.g. due to unequal access to SARS-CoV-2 testing or missing data. Whenever possible, outliers in our data were verified at source. We have not obtained the data about catecholamine treatment. The broad MIS-C case definition and advanced epidemiological situation allowed children with alternative diagnoses and coincidental positive SARS-CoV-2 results to be included in our cohort.

Precise epidemiological data on COVID-19 prevalence among age groups in Poland is lacking. We have also assumed homogeneity of risk of contracting the virus in the juvenile population.

## Conclusions

The severity of MIS-C is not as uniform as it seemed based on previous reports. In particular, ethnicity and sex may affect MIS-C phenotype. Consequently, management protocols might be not universally applicable, and should rather be adjusted to the specific population.

## Data Availability

Data on which manuscript is based is available on Mendeley Data under the CC-BY license.

http://dx.doi.org/10.17632/7nh8mrznkc.1

## Conflict of interest

none declared

## Acknowledgements

We thank to all the pediatricians who contributed to our work by reporting patients, especially to: Alicja Czajka, Tomasz Szatkowski, Marta Barszcz, Joanna Stryczyńska-Kazubska, Lidia Stopyra, Barbara Szczepańska, Marzena Zielinska, Katarzyna Mazur-Melewska, Sebastian Brzuszkiewicz, Ewelina Gowin, Bartosz Siewert, Robert Szylo, Magdalena Kosmider-Zurawska, Katarzyna Zieba-Glonek, Agnieszka Stroba-Żelek, Joanna Mańdziuk, Maciej Szczukocki, Justyna Kiepuszka, Katarzyna Rojewska, Elżbieta Berdej-Szczot, Ewa Czerwińska, Martyna Kukawska, Agnieszka Koczwara, Małgorzata Firek-Pedras, Katarzyna łapacz, Filip Tyc, Olga Izdebska, Danuta Koszałko, Anna Rożnowska-Wójtowicz, Agnieszka Maliszak, Paulina Opalińska-Zielonka, Paulina Sobiczewska, Anita Lackowska, Ewa Wołowska, Ilona Pałyga-Bysiecka, and to Aleksandra Tracewska for proofreading.

## Notes

### Competing Interest Statement

The authors have declared no competing interest.

### Clinical Trial

NCT04811456

### Funding Statement

Research was supported by a grant:
Research University-Excellence initiative
PSP: 501-D402-20-0006100

### Author Declarations

Ethical approval was obtained from the Bioethics Committee at Wroclaw Medical University (CWN UMW BW: 313/2020)

