## Supplement for "Multisystem inflammatory syndrome in European White children – study of 274 cases"

9)

**Supplementary Table 1. The Inclusion Criteria for MultiOrgan Inflammatory Syndromes COVID-19 Related Study (MOIS-CoR Study)**

| **Study inclusion criteria: age, disease severity, timing, diagnosis criterion and exclusion of other causes must be fulfilled.** |
|---|
| Age: 0-18 years |
| Disease severity: requiring hospitalization |
| Time frame: since March 4th 2020, ongoing (the paper covers data reported before 20th February 2021) |
| Diagnosis:<br>Kawasaki disease (KD) OR incomplete (atypical) Kawasaki disease (aKD)<br>OR toxic shock syndrome (TSS) OR macrophage activation syndrome[a] (MAS) OR unspecified inflammatory syndrome |
| **Kawasaki disease (KD) case definition[b]:**<br>Fever for at least 5 days and 4 from the following symptoms:<br>a) Erythema and cracking of lips, strawberry tongue, and/or erythema of oral and pharyngeal mucosa<br>b) Bilateral bulbar conjunctival injection without exudate<br>c) Rash: maculopapular, diffuse erythroderma, or erythema multiforme-like<br>d) Erythema and oedema of the hands and feet in acute phase and/or periungual desquamation in subacute phase<br>e) Cervical lymphadenopathy (≥1.5 cm diameter)<br><br>**Incomplete (atypical) Kawasaki disease (aKD) case definition:**<br>Fever for at least 5 days and 2 or 3 from the above symptoms OR infant with unexplained fever for at least 7 days AND CRP ≥3 mg/dL and/or ESR ≥40 mm/hr AND<br>1) at least 3 of the following:<br>a) Anaemia for age[c]<br>b) PLT ≥450 $\times 10^{9}$/L after the 7th day of fever<br>c) Albumin ≤3 g/dL<br>d) ALT ≥40 U/L<br>e) WBC count of ≥15 $\times 10^{9}$/L<br>f) Urine ≥10 WBC/hpf<br>OR<br>2) Changes in echocardiogram suggesting KD<br><br>**Toxic shock syndrome (TSS) case definition[d]:**<br>1) Fever AND<br>2) Hypotension[e] AND<br>3) at least two of the following organ systems involvement:<br>a) Gastrointestinal (vomiting, diarrhoea, abdominal pain);<br>b) Muscular (severe myalgia, elevated creatine phosphokinase level);<br>c) Renal (sterile pyuria, elevated creatinine or urea);<br>d) Hepatic (elevated liver enzymes and/or bilirubin level);<br>e) Hematologic (decrease in PLT $<100\times10^{9}$/L);<br>f) Disseminated intravascular coagulation;<br>g) Acute onset of diffuse pulmonary infiltrates and hypoxemia;<br>h) Acute onset of generalized oedema, or pleural or peritoneal effusions with hypoalbuminemia;<br>i) Central nervous system (alterations in consciousness in absence of fever and hypotension)<br><br>**Macrophage activation syndrome (MAS) case definition[f]:**<br>Febrile patient with:<br>1) Ferritin >684 ng/mL AND<br>2) Any two of the following:<br>a) PLT ≤181 $\times 10^{9}$/L<br>b) AST >48 U/L<br>c) Triglycerides >156 mg/dL<br>d) Fibrinogen ≤360 mg/dL<br><br>**Inflammatory syndrome case definition[g]:**<br>1) Fever for at least 3 days AND<br>2) High inflammatory markers (neutrophil count, CRP, ESR, procalcitonin) AND<br>3) Features of at least one organ dysfunction AND<br>4) At least two of the following symptoms:<br>a) Rash or bilateral non-purulent conjunctivitis or mucocutaneous inflammation signs (oral, hands or feet).<br>b) Hypotension or shock.<br>c) Features of myocardial dysfunction, pericarditis, valvulitis, or coronary abnormalities (including echocardiogram findings or elevated troponin/BNP/NT-proBNP),<br>d) Evidence of coagulopathy (by elevated INR, PTT, D-Dimers).<br>e) Acute gastrointestinal problems (diarrhoea, vomiting, or abdominal pain); |
| Exclusion of other infectious and non-infectious causes that could be responsible for the disease |

| |
|---|
| SARS-CoV-2 testing may be positive or negative. |

Abbreviations: ALT, alanine transaminase; AST, aspartate aminotransferase; BNP, brain natriuretic peptide; CRP, C-reactive protein; ESR, erythrocyte sedimentation rate; hpf, high power field; INR, international normalized ratio, NT-proBNP, N-terminal prohormone of brain natriuretic peptide; PLT, platelet count; PTT, partial thromboplastin time; SARS-CoV-2, severe acute respiratory syndrome coronavirus 2; WBC, white blood cells count

[a] DIC diagnosed using modified DIC score[1]
[b] Diagnostic criteria of KD and aKD in its typical and atypical (aKD) form were adapted from American Heart Association guidelines[2]
[c] Haemoglobin norms adapted from Cheng et al.[3]
[d] TSS was defined based on modified criteria by Centers of Disease Control and Prevention[4,5]
[e] Hypotension defined by a minimal systolic blood pressure (sBP) below 70+2×age (in years) mmHg or below 90 mmHg for children over 10 years old[6]
[f] MAS diagnosed using Paediatric Rheumatology International Trials Organization criteria[7]
[g] Adapted from the World Health Organization (WHO) multisystem inflammatory syndrome in children (MIS-C) definition[8]

**Supplementary Table 2. SARS-CoV-2 laboratory tests and anamnesis**

| | Cases | Percent of tested | Percent of all |
|---|---|---|---|
| **Contact with a confirmed COVID-19 case preceding MIS-C** | 120 | 51.9% | 43.8% |
| **Confirmed SARS-CoV-2 infection preceding MIS-C** | 24 | 9.8% | 8.8% |
| **Positive SARS-CoV-2 RT-PCR result at the time of MIS-C** | 29 | 12.6% | 10.6% |
| **Positive antibodies against SARS-CoV-2 at the time of MIS-C** | 241 | 94.5% | 88.0% |

Abbreviations: COVID-19, coronavirus disease 2019; MIS-C, multisystem inflammatory syndrome in children; RT-PCR, real-time transcription polymerase chain reaction; SARS-CoV-2, severe acute respiratory syndrome coronavirus 2

**Supplementary Table 3. Demographic and clinical characteristics, management and outcome of MIS-C cohort**

| Characteristic | Overall (N=274) | <5 y (N=64) | 5-12 y (N=140) | 12-18 y (N=70) | p | p gender-adjusted | PICU patients (N=23) | non-PICU patients (N=251) | p | p age-adjusted | p gender-adjusted |
|---|---|---|---|---|---|---|---|---|---|---|---|
| | n or med (% or IQR) | n or med (% or IQR) | n or med (% or IQR) | n or med (% or IQR) | | | n or med (% or IQR) | n or med (% or IQR) | | | |
| Male gender | 172 (63%) | 30 (47%) | 91 (65%) | 51 (73%) | <0.01 | 1 | 17 (74%) | 155 (62%) | 0.2 | 0.5 | 1 |
| Age (years) | 8.8 (5.2-12.1) | 3.2 (2.1-4.3) | 8.7 (6.6-10.5) | 13.6 (12.8-14.8) | <0.01 | <0.01 | 11.2 (10.1-12.6) | 8.4 (5.0-11.9) | <0.01 | 0.03 | <0.01 |
| BMI Z-score | 0.1 (-0.8-0.9) | -0.3 (-0.9-0.7) | 0.1 (-0.8-1.1) | 0.5 (-0.3-0.8) | 0.07 | 0.04 | 0.7 (-0.3-0.9) | 0.0 (-0.8-0.9) | 0.2 | 0.3 | 0.2 |
| **Comorbidities** | 38 (18%) | 7 (14%) | 21 (20%) | 10 (19%) | 0.7 | 0.7 | 4 (21%) | 34 (18%) | 0.7 | 0.8 | 0.7 |
| Obesity | 16 (7%) | 2 (4%) | 11 (9%) | 3 (5%) | 0.3 | 0.3 | 3 (14%) | 13 (6%) | 0.1 | 0.2 | 0.09 |
| Asthma | 11 (4%) | 2 (3%) | 6 (4%) | 3 (4%) | 0.9 | 0.9 | 1 (5%) | 10 (4%) | 0.9 | 1 | 0.9 |
| Chronic neurological issue | 6 (2%) | 2 (3%) | 3 (2%) | 1 (1%) | 0.8 | 0.7 | - | 6 (2%) | 0.5 | 0.5 | 0.4 |
| Other | 9 (4%) | 2 (4%) | 4 (3%) | 3 (6%) | 0.8 | 0.8 | - | 9 (4%) | 0.3 | 0.3 | 0.3 |
| **Symptoms and signs** | | | | | | | | | | | |
| **Fever length (days)** | 7.0 (6.0-9.0) | 7.0 (6.0-8.0) | 7.5 (6.0-9.0) | 7.0 (6.0-9.0) | 0.3 | 0.3 | 8.0 (7.0-10.0) | 7.0 (6.0-9.0) | 0.03 | .06 | 0.04 |
| **Mucocutaneous and lymph nodes** | 262 (97%) | 59 (95%) | 138 (99%) | 65 (94%) | 0.2 | 0.2 | 22 (96%) | 240 (97%) | 0.8 | 0.8 | 0.7 |
| Rash | 218 (83%) | 53 (84%) | 118 (87%) | 47 (71%) | 0.02 | 0.02 | 15 (71%) | 203 (84%) | 0.2 | 0.2 | 0.2 |
| Erythema at BCG site | 1 (0%) | 1 (2%) | - | - | 0.2 | 0.4 | - | 1 (0%) | 0.8 | 0.9 | 0.8 |
| Conjunctivitis | 207 (78%) | 47 (76%) | 109 (81%) | 51 (75%) | 0.6 | 0.6 | 14 (67%) | 193 (79%) | 0.2 | 0.2 | 0.2 |
| Hands/feet swelling or erythema | 142 (55%) | 39 (64%) | 75 (56%) | 28 (45%) | 0.1 | 0.2 | 15 (68%) | 127 (54%) | 0.2 | 0.1 | 0.1 |
| Oral inflammation | 173 (66%) | 43 (68%) | 96 (72%) | 34 (52%) | 0.02 | 0.02 | 14 (67%) | 159 (66%) | 0.9 | 0.8 | 1 |
| Cervical lymphadenopathy | 98 (38%) | 30 (48%) | 46 (36%) | 22 (32%) | 0.1 | 0.07 | 7 (33%) | 91 (38%) | 0.7 | 0.9 | 0.6 |
| **Gastrointestinal** | 250 (93%) | 56 (90%) | 132 (95%) | 62 (90%) | 0.3 | 0.3 | 21 (91%) | 229 (93%) | 0.8 | 0.8 | 0.6 |
| Abdominal pain | 222 (85%) | 42 (76%) | 120 (88%) | 60 (87%) | 0.1 | 0.2 | 21 (91%) | 201 (85%) | 0.4 | 0.5 | 0.5 |
| Nausea | 162 (62%) | 28 (47%) | 100 (75%) | 34 (51%) | <0.01 | <0.01 | 16 (70%) | 146 (61%) | 0.4 | 0.5 | 0.5 |
| Diarrhoea | 164 (62%) | 36 (59%) | 84 (61%) | 44 (65%) | 0.8 | 0.9 | 18 (78%) | 146 (60%) | 0.09 | 0.1 | 0.1 |
| **Upper respiratory** | 98 (38%) | 30 (54%) | 42 (31%) | 26 (39%) | 0.02 | 0.03 | 10 (45%) | 88 (38%) | 0.5 | 0.3 | 0.4 |
| Sore throat | 86 (34%) | 25 (47%) | 38 (28%) | 23 (35%) | 0.05 | 0.05 | 8 (36%) | 78 (34%) | 0.8 | 0.6 | 0.8 |
| Rhinitis | 23 (9%) | 10 (16%) | 7 (5%) | 6 (9%) | 0.05 | 0.09 | 2 (10%) | 21 (9%) | 0.9 | 0.7 | 0.8 |
| **Lower Respiratory** | 128 (50%) | 23 (40%) | 64 (48%) | 41 (63%) | 0.04 | 0.05 | 18 (86%) | 110 (47%) | <0.01 | <0.01 | <0.01 |
| Chest pain | 48 (19%) | 3 (6%) | 19 (14%) | 26 (39%) | <0.01 | <0.01 | 5 (25%) | 43 (18%) | 0.5 | 1 | 0.5 |
| Cough | 74 (28%) | 17 (28%) | 33 (25%) | 24 (35%) | 0.3 | 0.2 | 7 (32%) | 67 (28%) | 0.7 | 0.8 | 0.6 |

| | | | | | | | | | | | |
|---|---|---|---|---|---|---|---|---|---|---|---|
| Dyspnoea | 63 (24%) | 8 (13%) | 36 (27%) | 19 (28%) | 0.08 | 0.1 | 14 (67%) | 49 (20%) | <0.01 | <0.01 | <0.01 |
| **Neurological** | 220 (86%) | 54 (86%) | 112 (85%) | 54 (87%) | 0.9 | 0.9 | 20 (100%) | 200 (84%) | 0.06 | 0.06 | 0.05 |
| Lethargy | 151 (59%) | 36 (61%) | 88 (66%) | 27 (44%) | 0.02 | 0.02 | 19 (90%) | 132 (57%) | <0.01 | <0.01 | <0.01 |
| Irritability | 108 (42%) | 40 (65%) | 52 (39%) | 16 (25%) | <0.01 | <0.01 | 6 (27%) | 102 (43%) | 0.2 | 0.4 | 0.2 |
| Headache | 112 (46%) | 16 (32%) | 67 (51%) | 29 (47%) | 0.07 | 0.04 | 8 (38%) | 104 (47%) | 0.4 | 0.3 | 0.5 |
| Meningeal signs | 27 (10%) | 6 (10%) | 15 (11%) | 6 (9%) | 0.9 | 0.9 | 2 (9%) | 25 (11%) | 0.8 | 0.8 | 0.8 |
| Seizures | 3 (1%) | 2 (3%) | 1 (1%) | - | 0.2 | 0.06 | - | 3 (1%) | 0.6 | 0.7 | 0.6 |
| Muscle hypotension | 30 (11%) | 5 (8%) | 16 (12%) | 9 (14%) | 0.6 | 0.7 | 5 (24%) | 25 (10%) | 0.07 | 0.1 | 0.07 |
| Nerve paralysis | 2 (1%) | 1 (2%) | 1 (1%) | - | 0.6 | 0.6 | - | 2 (1%) | 0.7 | 0.7 | 0.7 |
| Nerve paresis | 2 (1%) | - | 2 (1%) | - | 0.4 | 0.4 | - | 2 (1%) | 0.7 | 0.7 | 0.7 |
| Smell loss | 8 (3%) | 1 (2%) | 2 (1%) | 5 (8%) | 0.05 | 0.02 | 1 (5%) | 7 (3%) | 0.7 | 0.9 | 0.6 |
| Taste loss | 7 (3%) | 1 (2%) | 2 (1%) | 4 (6%) | 0.2 | 0.08 | 1 (5%) | 6 (3%) | 0.6 | 0.7 | 0.5 |
| Photophobia | 28 (11%) | 8 (14%) | 16 (12%) | 4 (6%) | 0.4 | 0.4 | - | 28 (12%) | 0.1 | 0.1 | 0.1 |
| Skin hyperesthesia | 87 (34%) | 29 (48%) | 46 (35%) | 12 (19%) | <0.01 | <0.01 | 8 (36%) | 79 (34%) | 0.8 | 0.4 | 0.7 |
| **Cardiovascular** | 139 (58%) | 16 (33%) | 79 (63%) | 44 (69%) | <0.01 | <0.01 | 22 (100%) | 117 (54%) | <0.01 | <0.01 | <0.01 |
| Hypotension[a] | 99 (41%) | 7 (14%) | 57 (46%) | 35 (51%) | <0.01 | <0.01 | 18 (86%) | 81 (36%) | <0.01 | <0.01 | <0.01 |
| Coronary dilation or aneurysm[b] | 21 (8%) | 6 (11%) | 10 (7%) | 5 (8%) | 0.8 | 0.5 | 3 (14%) | 18 (8%) | 0.3 | 0.3 | 0.4 |
| EF<55% | 58 (23%) | 3 (5%) | 31 (23%) | 24 (39%) | <0.01 | <0.01 | 14 (64%) | 44 (19%) | <0.01 | <0.01 | <0.01 |
| Pericardial effusion | 24 (9%) | 5 (9%) | 15 (11%) | 4 (6%) | 0.6 | 0.5 | 3 (14%) | 21 (9%) | 0.5 | 0.5 | 0.6 |
| **Musculo-osteoarticular** | 111 (44%) | 17 (32%) | 58 (44%) | 36 (55%) | 0.04 | 0.03 | 8 (42%) | 103 (44%) | 0.8 | 0.6 | 0.9 |
| Muscle pain | 103 (41%) | 15 (29%) | 53 (40%) | 35 (54%) | 0.02 | 0.01 | 8 (42%) | 95 (41%) | 0.9 | 0.8 | 0.9 |
| Arthralgia | 50 (19%) | 6 (10%) | 29 (21%) | 15 (24%) | 0.1 | 0.08 | 4 (20%) | 46 (19%) | 0.9 | 0.8 | 0.9 |
| Arthritis | 12 (5%) | 1 (2%) | 6 (4%) | 5 (8%) | 0.2 | 0.2 | 1 (5%) | 11 (5%) | 1 | 0.8 | 1 |
| Scrotum/labia swelling | 26 (10%) | 8 (13%) | 12 (9%) | 6 (10%) | 0.6 | 0.2 | 3 (14%) | 23 (10%) | 0.5 | 0.4 | 0.7 |
| **Clinical presentation** | | | | | | | | | | | |
| KD[c] | 170 (67%) | 43 (70%) | 94 (71%) | 33 (54%) | 0.06 | 0.09 | 18 (86%) | 152 (65%) | 0.05 | 0.03 | 0.04 |
| MAS[d] | 59 (22%) | 4 (6%) | 30 (22%) | 25 (36%) | <0.01 | <0.01 | 9 (43%) | 50 (20%) | 0.02 | 0.08 | 0.02 |
| DIC[e] | 42 (17%) | 5 (9%) | 22 (17%) | 15 (22%) | 0.1 | 0.06 | 5 (22%) | 37 (16%) | 0.5 | 0.7 | 0.4 |
| **Therapy and outcome** | | | | | | | | | | | |
| Admission since onset (days) | 5.0 (4.0-6.0) | 5.0 (4.0-5.0) | 5.0 (4.0-6.0) | 5.0 (4.0-6.5) | 0.5 | 0.5 | 6.0 (4.0-7.0) | 5.0 (4.0-6.0) | 0.08 | 0.1 | 0.09 |
| Intensive care | 23 (8%) | 1 (2%) | 13 (9%) | 9 (13%) | 0.05 | 0.08 | 23 (100%) | - | <0.01 | <0.01 | <0.01 |
| Mechanical ventilation | 10 (4%) | - | 7 (5%) | 3 (5%) | 0.2 | 0.2 | 10 (43%) | - | <0.01 | <0.01 | <0.01 |
| Oxygen supplementation | 61 (23%) | 5 (8%) | 41 (31%) | 15 (23%) | <0.01 | <0.01 | 17 (77%) | 44 (18%) | <0.01 | <0.01 | <0.01 |

| | | | | | | | | | | | |
|---|---|---|---|---|---|---|---|---|---|---|---|
| No immunomodulatory agent | 8 (3%) | 3 (5%) | 3 (2%) | 2 (3%) | 0.6 | 0.4 | - | 8 (3%) | 0.4 | 0.4 | 0.3 |
| IVIG | 238 (93%) | 55 (90%) | 122 (95%) | 61 (91%) | 0.3 | 0.3 | 20 (100%) | 218 (92%) | 0.2 | 0.2 | 0.2 |
| GCS | 143 (67%) | 27 (56%) | 74 (69%) | 42 (71%) | 0.2 | 0.2 | 17 (89%) | 126 (64%) | 0.03 | 0.04 | 0.03 |
| IVIG and GCS | 133 (62%) | 24 (50%) | 69 (65%) | 40 (67%) | 0.1 | 0.2 | 16 (89%) | 117 (60%) | 0.01 | 0.03 | 0.02 |
| Other immunomodulatory agents | 3 (1%) | 1 (2%) | 2 (2%) | - | 0.6 | 0.7 | - | 3 (2%) | 0.6 | 0.7 | 0.6 |
| ASA | 226 (85%) | 51 (81%) | 119 (88%) | 56 (84%) | 0.4 | 0.4 | 18 (90%) | 208 (85%) | 0.5 | 0.6 | 0.5 |
| Anticoagulation | 13 (5%) | 3 (5%) | 5 (4%) | 5 (7%) | 0.5 | 0.5 | 1 (5%) | 12 (5%) | 1 | 1 | 0.9 |
| Complete recovery at discharge | 218 (93%) | 55 (95%) | 115 (94%) | 48 (87%) | 0.2 | 0.2 | 18 (86%) | 200 (93%) | 0.2 | 0.3 | 0.2 |

Abbreviations: ASA, acetylsalicylic acid; BCG, Bacillus Calmette–Guérin vaccine; BMI, body mass index; DIC, disseminated intravascular coagulation; EF, ejection fraction; GCS, glucocorticoids; IQR, interquartile range; IVIG, intravenous immunoglobulin; KD, Kawasaki disease; MAS, macrophage activation syndrome; med, median; MIS-C, multisystem inflammatory syndrome in children; PICU, paediatric intensive care unit; y, years old

[a] Defined by a minimal systolic blood pressure below 70+2×age (in years) mmHg or below 90 mmHg for children over 10 years old[6]

[b] Dilation was defined as a Z-score between 2 to 2.5, while aneurysm as Z-score ≥2.5[2,9,10]

[c] Diagnostic criteria of KD in its typical and atypical (aKD) form were adapted from American Heart Association guidelines[2]

[d] MAS was diagnosed based on Paediatric Rheumatology International Trials Organization criteria[7]

[e] DIC was diagnosed using modified DIC score[1]

**Supplementary Figure 1. Geospatial distribution of MIS-C cases and reporting cities in Poland**

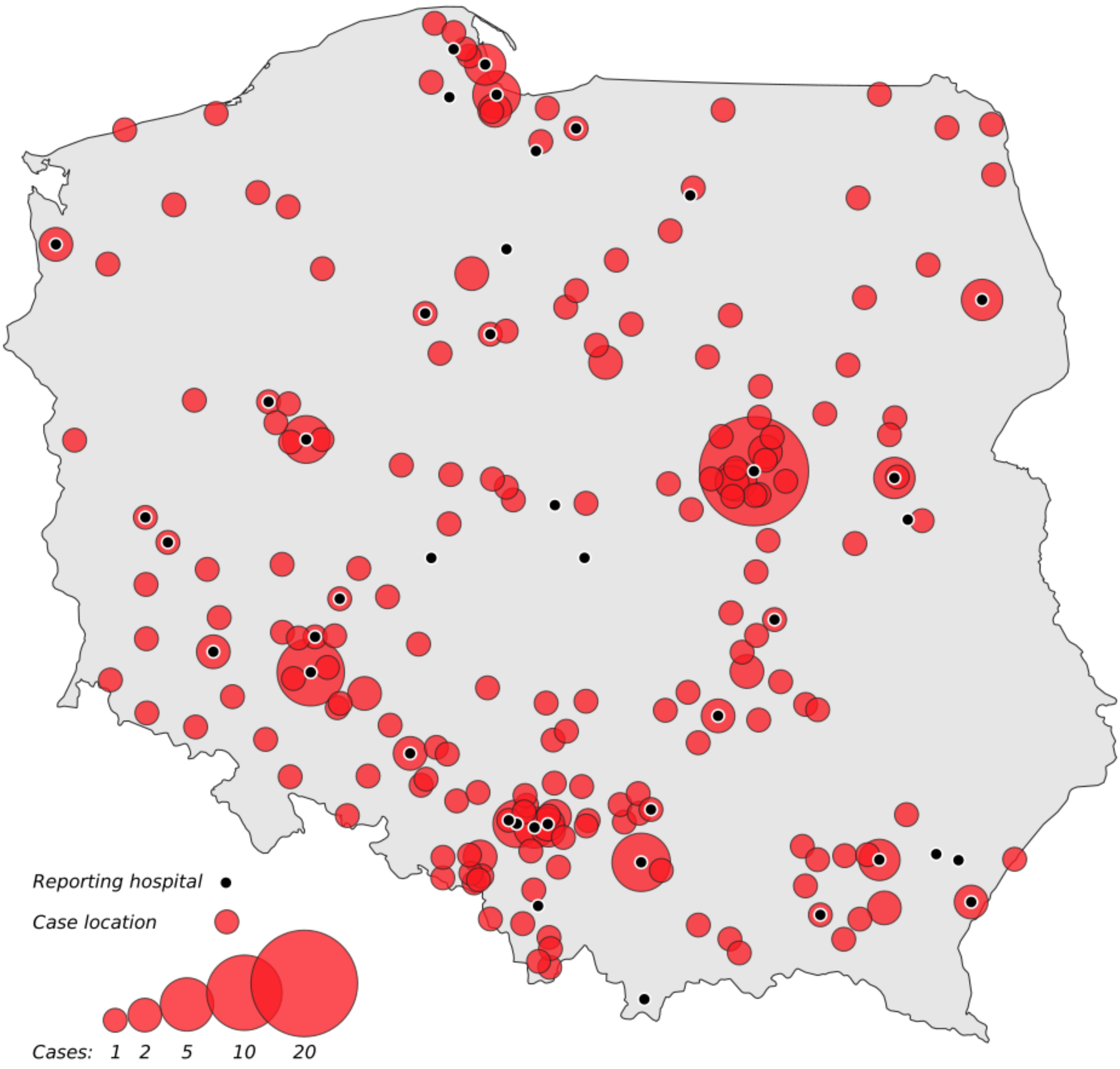

MIS-C, multisystem inflammatory syndrome in children
Registry data was based on voluntary case reporting by participating sentinel surveillance sites. Between 4th March 2020 (when the first COVID-19 case was confirmed in Poland) and by 20th February 2021 274 cases meeting MIS-C criteria were captured. 267 of 274 cases, for which location was known, are presented on the map above. MIS-C criteria were based on the World Health Organization definition (consult Materials and Methods section in the main text).
